# Targeting *de novo* loss of function variants in constrained disease genes improves diagnostic rates in the 100,000 Genomes Project

**DOI:** 10.1101/2022.05.18.22275260

**Authors:** Eleanor G. Seaby, N. Simon Thomas, Amy Webb, Helen Brittain, Ana Lisa Taylor Tavares, Genomics England Consortium, Diana Baralle, Heidi L. Rehm, Anne O’Donnell-Luria, Sarah Ennis

**Author notes:** <u>Corresponding author:</u> Dr Eleanor Seaby, Genomic Informatics Group, MP 808, Duthie Building, University Hospital Southampton, SO16 6YD. These authors contributed equally to this work (Anne O’Donnell-Luria, Sarah Ennis). **Ethics approval and consent to participate** All patients included in this study consented to participate in the 100,000 Genomes Project - ethics approval by the Health Research Authority (NRES Committee East of England) REC: 14/EE/1112; IRAS: 166046. The ethical approval letter is available upon request.

## Abstract

Whole genome sequencing was first offered clinically in the UK through the 100,000 Genomes Project (100KGP); however, data analysis was time and resource intensive with 3 million variants found per patient. Consequently, analysis was restricted to predefined gene panels associated with the patient’s phenotype. However, panels rely on clearly characterised phenotypes and risk missing diagnostic variants outside of the panel(s) applied. We propose a complementary method to rapidly identify diagnostic variants, including those missed by 100KGP methods.

The Loss-of-function Observed/Expected Upper-bound Fraction (LOEUF) score quantifies gene constraint, with low scores correlated with haploinsufficiency. We applied DeNovoLOEUF, a filtering strategy to sequencing data from 13,949 rare disease trios in the 100KGP, by filtering for rare, *de novo*, single nucleotide loss-of-function variants in OMIM disease genes with a LOEUF score <0.2. We conducted our analysis prospectively in 2019 and compared our findings with the corresponding diagnostic reports as returned in 2019 and again in 2021.

324/336 (96%) of the variants identified through DeNovoLOEUF were classified as diagnostic or partially diagnostic. We identified 39 diagnoses that were “missed” by 100KGP standard analyses, which are now being returned to patients. We have demonstrated a highly specific and rapid method with a 96% positive predictive value that has good concordance with standard analysis, low false positive rate, and can identify additional diagnoses. Globally, as more patients are being offered genome sequencing, we anticipate that DeNovoLOEUF will rapidly identify new diagnoses and facilitate iterative analyses when new disease genes are discovered.

## Introduction

With transformative advances in genomic medicine, there has been an exponential rise in the number of individuals undergoing exome and whole genome sequencing. A shift towards large-scale international sequencing programs is improving affordability and accessibility of such sequencing for diagnostic purposes, where conventional clinical tests have failed to yield a diagnosis.^1,2^ The 100,000 Genomes Project (100KGP), was a research project embedded within the UK National Health Service and the precursor to offering whole genome sequencing (WGS) as a clinical test.^3-5^ This pioneering project benefited from sequencing vast patient numbers with rare genetic diseases with improved power to identify multiple patients with overlapping phenotypes and genotypes, however the number of cases that required clinical assessment for diagnostic reporting versus resources available created a significant bottleneck.

Diagnostic rates for the 100KGP were similar to the international average for rare diseases.^6^ The flagship 100KGP paper showed that an estimated diagnostic uplift from 15% to 20% could be achieved beyond prior testing but that the time and level of additional resources required to analyse the full genome was beyond routine diagnostic testing.^5^ As a result, the project adopted the use of predefined gene panels (Genomics England PanelApp)^7^ to target sequencing analysis to the most relevant genes selected from the Human Phenotype Ontology (HPO) terms provided by referring clinician (**Figure 1**).^5^ Whilst this approach restricted the number of variants assessed and improved the efficiency of the variant curation process applied to each patient’s genome for diagnostic reporting, it risked missing variants in genes outside of the gene panel applied. At the latter stages of the project, many NHS England clinical laboratories additionally reviewed all *de novo* variants and top Exomiser^8^ results, although this was never mandatory. This learning has also informed guidance for the evaluation of whole genome sequencing, which is now available as a clinical test in the NHS.

**Figure 1.**
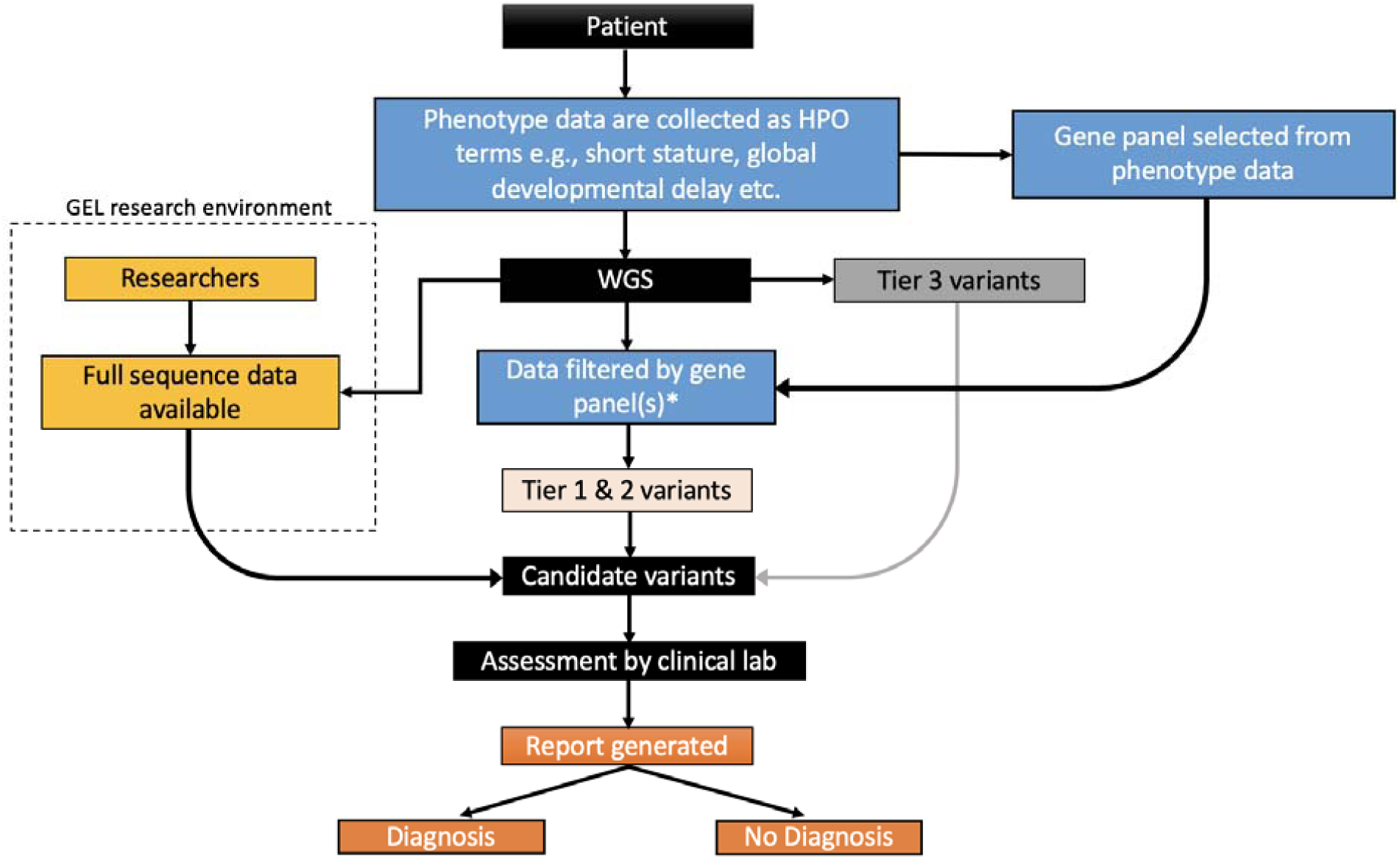
Genomics England 100,000 Genomes Project workflow Phenotype data were collected from patients and recorded as HPO terms. These terms informed the virtual gene panel(s) applied for data analysis. This contrasts with the new UK Genome Medicine Service, where the clinician selects the panel(s) applied. In 100KGP, the patient underwent whole genome sequencing (WGS), and their sequencing data were filtered using the pre-selected gene panel(s). Data were also filtered by allele frequency and variant segregation (*) and these variants were classified into Tier 1 and Tier 2 variants as previously described.^5^ Candidate variants within the gene panel were identified and assessed by an NHS accredited diagnostic laboratory and a report was generated and returned to the patient. Diagnostic laboratories were under no obligation to review variants outside of the virtual panel(s) applied, however rare variants outside of the predefined panel(s) were available for analysis as Tier 3 variants. Variants outside of the gene panel(s), including full raw sequencing data, remain accessible to approved researchers for interrogation. Potential candidate variants identified through this route can be reported via Genomics England for potential return for local review by clinical laboratories.

Accurate phenotyping is essential for gene panel selection, yet there is huge variability in the phenotypes reported by clinicians. For some cases in the 100KGP, only a single HPO term was reported. As WGS becomes more widespread, appropriate matching of HPO terms with optimal panel(s) may be less error prone for experienced geneticists but will represent a challenge for the wider community of clinicians expected to routinely refer patients.

Furthermore, whilst the use of HPO terms aids in the standardization of reporting phenotype data, it represents a cross-sectional timepoint analysis without resource for reanalysis. Ultimately, using HPO terms and relying on predefined gene panel options lacks the full clinical narrative and challenges gene panel selection. Therefore, there is need to expand genome analysis beyond gene panels to enable a more agnostic and comprehensive whole genome analysis, yet this needs to be balanced with the number of variants that require manual assessment for diagnostic reporting. Targeting variants with high pathogenic potential across the entire exome provides an opportunity to rapidly identify diagnostic variants and uplift diagnostic rates. Genotype-driven analysis approaches are complementary to the phenotype-drive approach currently utilized by 100KGP.

Looking at variation across people, some genes are extremely depleted or constrained for variation predicted to result in loss-of-function (LoF).^9^ That is to say there is negative selection against the loss or inactivation of one allele. By comparing the observed over the expected rate of predicted loss-of-function variants in large population databases, it is now possible to compute the degree of constraint a given gene has for inactivation.^10,11^ The loss-of-function observed over expected upper bound fraction, or LOEUF score, is a metric that places each gene on a continuous scale of loss-of-function constraint. Low scores are highly correlated with disease genes and gene essentiality, with the first LOEUF decile being enriched for haploinsufficient disease genes and the greatest burden of OMIM disease entries.^11^ Loss of function variants in extremely LoF constrained genes are therefore prime targets for potential diagnoses.

We aimed to utilise the sequencing and phenotype data generated through 100KGP and apply a transferable and rapid filtering method, we called DeNovoLOEUF, that can screen for putative pathogenic variants. We developed a rapid, agnostic approach to target the highest diagnostic yield variants in rare disease patients whilst enabling clinical curators to focus on the most important findings, regardless of the gene panel applied, improving efficiency for cases where a diagnosis could be rapidly identified.

## Subjects and Methods

### Data access

We obtained access to the GEL research environment (RE) and high-performance cluster (HPC) behind a secure firewall following information governance training and with membership of a Genomics England Clinical Interpretation Partnership (GeCIP): *Quantitative methods, machine learning, and functional genomics*. We had a project approved (RR359 - *Translational genomics: Optimising novel gene discovery for 100,000 rare disease patients*) which permitted access to 100KGP sequencing and phenotype data. This included an aggregate .*gvcf* file comprising 13,949 rare disease trios with *de novo* variants, called using the Illumina Platypus pipeline.

### Phenotype data

Referring clinicians recorded phenotype data as discrete HPO terms. These were accessible in the RE by querying HPO terms stored in mysql tables in a LabKey data management system.

### Data analysis

Data analysis was performed in Autumn 2019 (**Figure 2**). Bespoke scripts were developed to query the aggregate .*gvcf* file. We selected only variants that passed Illumina QC as previously described.^5^ We then applied a filtering strategy called DeNovoLOEUF: Firstly, we selected *de novo* variants with an allele frequency <0.001 in gnomAD v2.1.1 (all populations). We further restricted this list to predicted loss of function (pLoF) variants including nonsense, frameshift and essential (canonical) splice site variants. We imported LOEUF constraint gene scores, downloaded from the gnomAD browser (https://gnomad.broadinstitute.org), into the research environment. We then restricted rare, *de novo*, pLoF variants to genes with a LOEUF score of <0.2 (n=1044), approximately equivalent to the first LOEUF decile, representing a list of genes most highly constrained for loss-of-function and predicted to cause disease through haploinsufficiency. We retained only those LOEUF constrained genes with known disease gene association in the OMIM^12^ database (n=335), accessed and downloaded as a flat .*txt* file in October 2019. Variants in novel disease genes represent further potential diagnoses but are beyond the scope of this disease-gene focused assessment.^13^ We applied LOFTEE v1.0^11^ to flag variants as potential false positives but retained variants in the terminal exon. Variants remaining following DeNovoLOEUF filtering steps were considered *putative diagnostic variants*.

**Figure 2.**
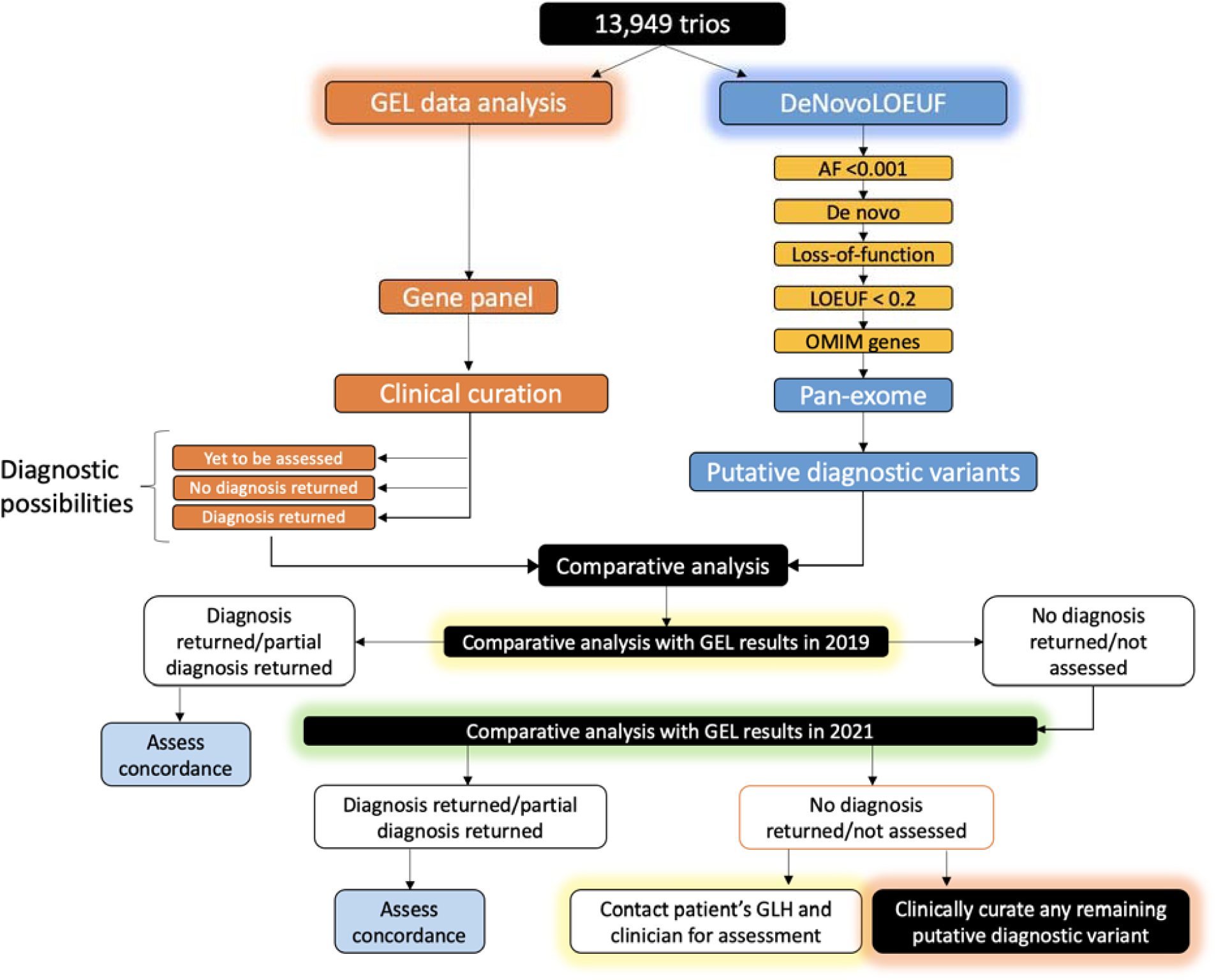
Summary of methods Putative diagnostic variants identified by our filtering approach were compared with the diagnostic reports for the same patients. Following comparative analysis in 2021, if a negative report had been issued, or the case was still under review, we contacted the patient’s Genomics Laboratory Hub and referring clinician and shared our variant of interest. If we received no response, we clinically curated the remaining variants as per ACMG-AMP guidelines in an NHS approved diagnostic laboratory. AF – allele frequency; GEL – Genomics England; GLH – Genomics Laboratory Hub; LOEUF – Lower Observed/Expected Upper-bound Fraction; OMIM – Online Mendelian Inheritance in Man.

Clinical outcome data pertaining to diagnostic reports and individual specific phenotype information were extracted querying Labkey using the RLabKey package in R. These phenotype data were merged with filtered putative pathogenic variants for each patient. We further extracted the diagnostic report status for each patient, which included any returned pathogenic variants, by querying the ‘GMC exit questionnaire’ table in LabKey.

### Comparative analysis

Putative diagnostic variants were compared with the diagnoses returned to patients recruited to 100KGP at two time points (October 2019 and April 2021) to assess concordance between our DeNovoLOEUF filtering method and the analysis strategy by GEL.

At the first time point, putative diagnostic variants extracted using DeNovoLOEUF were correlated against variants declared in the Genome Medicine Centre exit questionnaire of the RE as being returned to patients in their diagnostic report. This was to assess the positive predictive value of the method. It was expected that many patients would not have had a diagnostic report returned in 2019, i.e. their case status was “yet to be assessed”. The comparative analysis was therefore repeated at the second time point (18 months later in April 2021), to assess whether our method correctly predicted additional diagnoses determined over time as the proportion of closed 100KGP cases increased.

Cases that were not assessed or reported as negative (i.e. no diagnosis identified) by 2021, were re-curated by NHS Clinical Scientists to standardize curation of any novel diagnoses not originally detected through the 100KGP. This was achieved in two ways. Firstly, we contacted the patient’s Genomic Laboratory Hub (GLH), previously known as the Genome Medicine Centre, and referring clinician to discuss the variant we had found and asked whether the variant was already known about and/or had been returned as a diagnosis. Communication with the GLH and referring clinicians often prompted local multidisciplinary team meetings followed by diagnostic laboratory confirmation of the variant. Secondly, for the variants that were unknown to the GLHs, or for which we received no response from the centres contacted, we worked with clinical scientists in the Wessex Regional Genetics Laboratory, an established GEL diagnostic reporting centre, who curated the remaining variants alongside the patients phenotypes as per the ACMG-AMP guidelines.^14^ We then determined how many of the remaining putative diagnostics variants would meet a partial or full diagnosis.

## Results

A total of 383 putative diagnostic variants were identified by DeNovoLOEUF in 375 patients. Of these variants, 339/383 (88%) were in the Exomiser top ranked results. There were more variants than patients due to some individuals harbouring more than one *de novo* variant in the same gene (n_patients_ = 6) or having more than one *de novo* variant in two different genes (n_patients_ = 2). Results stratified by time-point assessment are shown in **Figure 3**.

**Figure 3.**
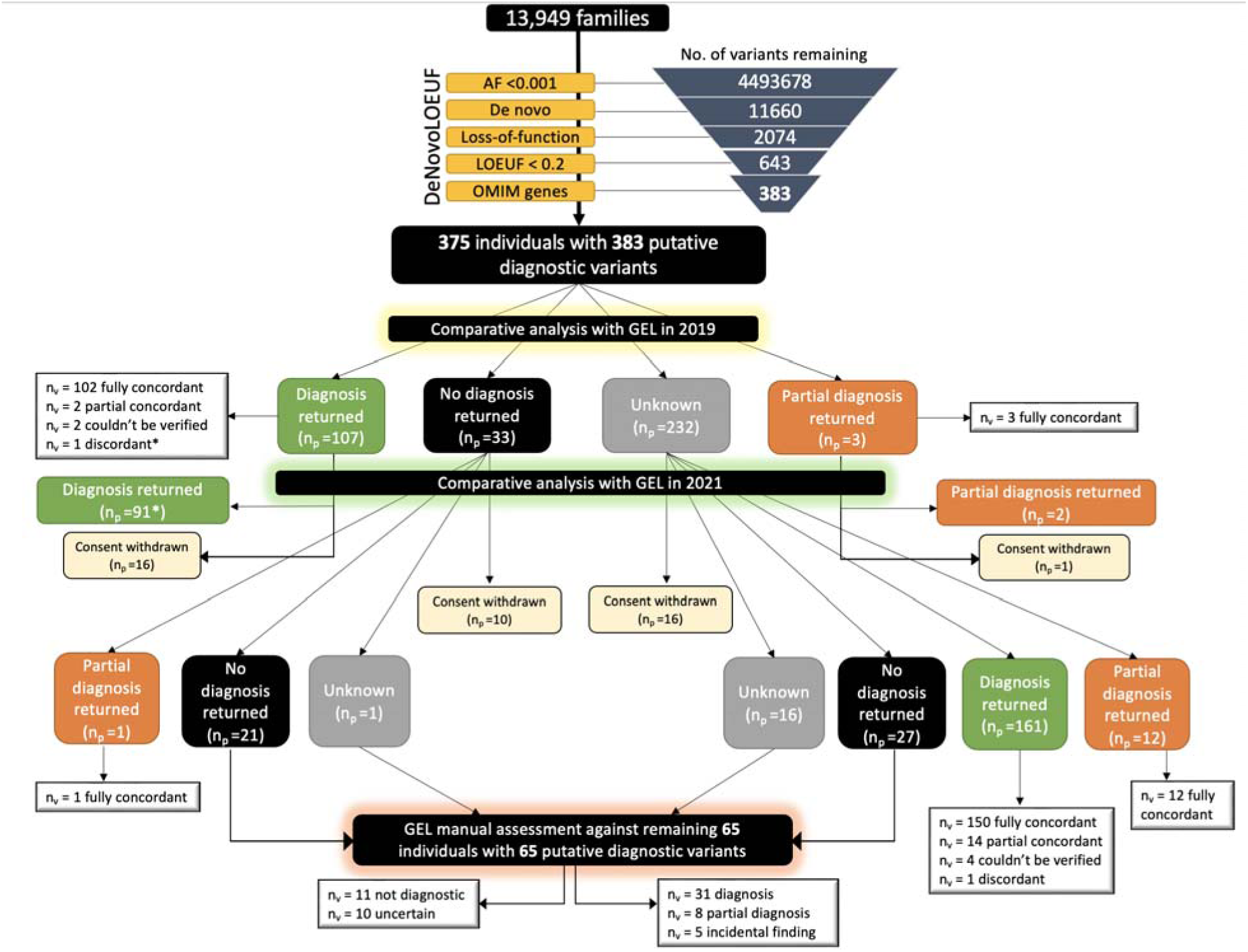
Summary of results using DeNovoLOEUF on 100,000 Genomes Project Patients In October 2019, 383 predicted loss of function variants in 375 individuals were identified in known OMIM disease genes using DeNovoLOEUF. At the time, 29% (107/375) of patients had a single diagnostic variant returned and 33 patients had their case closed as ‘negative’. Ninety-five percent (102/107) of variants identified by our method were entirely concordant with the formal GEL diagnosis returned. In two patients where we correctly identified a pathogenic variant, a second variant in a different gene also contributed to the diagnosis (partial concordant). Two variants were excluded from comparative analysis as it was not possible to verify the returned diagnosis in the GEL research environment’s exit questionnaire. One variant was discordant (*) between the variant identified by DeNovoLOEUF and the reported outcome data from the 100KGP; however, this variant was subsequently confirmed as pathogenic by 2021. Three patients had a partial diagnosis returned, meaning a single variant was returned to the patient but that it did not fully explain the phenotype; all three were fully concordant with our method. 232 cases were ‘unknown’ meaning that no formal report had been returned to the patient. In summary, at the first time point, the method correctly detected 99% (107/108) of reported diagnostic variants. For the comparative analysis in 2021, 43 variants were excluded from downstream analysis due to patients being withdrawn from 100KGP. A further 150 variants were fully concordant with our method, 14 were partially concordant, 13 variants were concordant with a returned partial diagnosis, and 1 variant was discordant. Following assessment of the remaining 65 variants in 65 individuals, 31 cases were considered diagnostic, 8 cases partially diagnostic, and 5 cases were incidental findings. Eleven variants did not explain the phenotype, and 10 cases were uncertain meaning that there was insufficient clinical information to determine causality. n_p_ = number of patients; n_v_ = number of variants.

### Comparative analysis of method in 2021

In April 2021, 284/383 (74%) of all variants initially identified were confirmed as either fully diagnostic or partially diagnostic, including 17 patients who had diagnostic reports returned in 2019 prior to being withdrawn from the study. A single discordant variant identified in 2019 (**Figure 3**) was reclassified as a pathogenic variant by 2021 and returned to the patient as diagnostic. Twenty-six patients harboring 26 variants identified in 2019 were unavailable for further assessment in 2021 due to either withdrawing from the 100KGP or data being temporarily removed from the trusted research environment whilst re-consent was sought for child participants reaching adulthood. Only one variant that we identified did not match the variant returned to the patient, meaning GEL had returned an alternative diagnosis, however the variant we identified is a known pathogenic variant in ClinVar. This patient had only one HPO term recorded “cystinosis”, which was consistent with the biallelic variants reported by GEL. We are attempting to contact the clinician responsible for this patient to gain further clinical information. Six variants could not be assessed for concordance as the variant returned by 100KGP was not identifiable in the GEL research environment. Sixty-five variants in 65 individuals identified using DeNovoLOEUF remained unresolved in 2021 and required further scrutiny. All 65 variants were in Tier 3 of the GEL tiering system as they were *de novo* loss-of-function variants.

### Assessment of remaining 65 variants

Seventeen (26%) of the remaining 65 variants identified by DeNovoLOEUF were confirmed as full diagnoses after discussion with the patient’s local GLH or referring clinician, prompting independent validation in local NHS laboratories. The remaining 48 genes were manually curated (**Supplementary Data**) by two clinical scientists working in an NHS accredited genomic diagnostic laboratory. Of these variants, 14 were designated diagnostic, 8 were partially diagnostic, and 5 were identified as incidental findings. Eleven variants were considered not diagnostic, and 10 cases were uncertain. Half (5/10) of the uncertain variants were classified as pathogenic but there was insufficient clinical information to confirm causality for the patient’s phenotype; the remainder were classified as variants of uncertain significance (VUSes). Of the 11 non-diagnostic cases, three were heterozygous variants in autosomal recessive disease genes with no second hit. One variant was in an X-linked recessive gene and the patient was female. A further variant passed GEL quality control filtering in 2019 but upon more detailed inspection was artefactual. Three variants were a poor phenotypic fit, and two variants were intronic on the Matched Annotation from NCBI and EBML-EBI (MANE)^15^ Select transcript and not present in an exon of a MANE Plus Clinical alternative transcript harboring known pathogenic variants.

### Summary of all results

In summary, 324/336 (96%) of the variants identified through DeNovoLOEUF filtering (excluding incidental findings, variants for withdrawn participants, or in patients where there was inadequate phenotype or genotype reporting in GEL) were classified as diagnostic or partially diagnostic (**Table 1**).

**Table 1.** Tabulated summary of results Summary of results comparing variants identified by our method compared with diagnostic reports returned to patients, in addition to variant curation. The 2021 comparison is a cumulative comparison of the 2021 and 2019 comparisons. Full concordance is where a single variant we identified was confirmed as the pathogenic variant. Partial concordance denotes the case where the variant we identified was pathogenic but did not explain the full phenotype, or a second variant in a different gene was returned to the patient. Variants that could not be verified were those where it was not possible to see which variant had been returned to the patient as diagnostic, or there was not enough clinical information to determine causality. These 16 variants, plus the 5 incidental findings were not included in the PPV calculations. GLH – Genomics Laboratory Hub; PPV – positive predictive value.

| GEL diagnoses returned | 2019 comparison | 2021 comparison | Following curation/GLH |
| --- | --- | --- | --- |
| Fully concordant | 102 | 253 | 284 |
| Partially concordant | 5 | 32 | 40 |
| Couldn't be verified | 2 | 6 | 16 |
| Discordant | 1 [discarded in 2021] | 1 | 12 |
| Incidental | 0 | 0 | 5 |
| PPV | 107/108 = 99% | 285/286 = 99% | 324/336 = **96%** |

## Discussion

We describe a fast, unbiased filtering strategy, DeNovoLOEUF, to identify potential pathogenic variants with high positive predictive value and specificity. We utilise the LOEUF spectrum of constraint to identify *de novo* loss of function variants with a high potential of pathogenicity. Unlike the approach adopted in the 100KGP with panel-based tiering, our genotype-driven method is agnostic to phenotype and independent of gene panels which often change over time and require the correct one to be selected. Indeed, we identified 39 diagnostic or partially diagnostic variants in 39 known disease-associated genes that had been missed by standard 100KGP diagnostic protocols due to the gene in which the causal variant was identified not being included on the gene panel selected, although 35 genes were included on different gene panels. In view of the recognition of diagnoses outside of the selected gene panel(s), some NHS accredited laboratories adopted a policy, when reviewing results from the 100KGP available within the 100K results portal, to assess *de novo* variation and Exomiser top-ranked results to uplift the resultant diagnoses. This approach to reporting has been informative for the strategy within subsequent large-scale WGS endeavors. When including these results, 26% of all causal variants returned by NHS labs in the 100KGP were not in the initial gene panel applied, exemplifying the issue with gene panel analysis strategies.^6^ However, re-analyses involve re-visiting sequencing data and clinical cases, something which could be mitigated by screening and prioritising highly putative diagnostic variants in the first instance. Using DeNovoLOEUF as a screening strategy would have immediately identified 324 pathogenic variants, saving considerable time and money. On average, DeNovoLOEUF added only one extra variant for assessment in ∼3% of all rare disease probands (0.023 variants per person).

Our method is rapid, having identified 175 variants in 2019 that we were unable to efficiently return to the patient’s clinical teams. As a result of our collaboration with colleagues at Genomics England, a form that enables the submission of multiple potential diagnoses for different participants via a single submission within the RE, is now in place. DeNovoLOEUF utilises an effective screening approach to detect highly penetrant putative diagnostic variants across a large cohort. It should however be noted that whilst 285/336 (85%) of all the variants identified were fully diagnostic, 40/336 (12%) were a partial diagnosis, meaning that the variant was considered pathogenic but did not fully explain the phenotype. Additionally, following manual curation there were 10 variants whereby we were unable to confirm whether the variant explained the phenotype; all these patients lacked sufficient clinical data to determine causality, even though five of the variants were pathogenic by ACMG-AMP guidelines. This highlights some of the challenges with phenotyping in a large-scale national sequencing project and that using HPO terms are sometimes insufficient to make a diagnosis and post-analysis communication with the clinical care team is a critical component of molecular diagnosis as emphasized by ACMG clinical practice guidelines.^16^ Genomics England is actively supporting improvements at the clinical-research interface to enable collaborations between researchers and clinicians and in patient phenotyping by the provision of Hospital Episode Statistics data within the RE as a longitudinal record of participants’ phenotypes.

With ever increasing application of genome sequencing and a drive to sequence a further 5 million genomes in the UK, there is clear demand to find efficient analytic strategies. We attest our method to be a suitable adjunct to the current protocols to identify causal variation in 100KGP, the NHS Genomics Medicine Service, or other similar international initiatives. DeNovoLOEUF is capable of prioritising putative pathogenic variants for diagnostic laboratories, saving time and resources. Furthermore, one of the draw backs of applying gene panels is that many are already outdated at the point of use, with new genes being consistently added to the literature. Our method can be easily applied iteratively for re-analysis as new genes are discovered but would benefit from including additional disease genes in ClinVar^17^, GenCC^18^, or HGMDPro that have yet to be indexed in OMIM.

Whilst the DeNovoLOEUF method has a high PPV, 12 variants (1 from the 2021 analysis and 11 from curation analysis) were discordant with the diagnosis returned. Four of the variants were in LOEUF constrained genes, but the genes caused disease through biallelic inheritance. We selected a LOEUF cut off <0.2 which is highly enriched for autosomal dominant disease genes, although this does not exclude a small number of recessive diseases.^11^ Our method might be further refined to assess less constrained genes, including additional haploinsufficient genes with LOUEF >0.2 through genes known to cause disease through recessive inheritance. It could also be expanded to evaluate other *de novo* variants in these genes with high *in silico* scores such as REVEL^19^ for missense variants and SpliceAI^20^ for variants that may impact splicing.

Two variants were pLoF on a non-canonical transcript that was poorly expressed across tissues,^21^ and intronic on the MANE Select transcript. Whilst one option would be to limit our method to variants on the MANE Select and MANE Plus Clinical transcripts, this is potentially problematic not all genes have been curated to define additional transcripts to be included in the MANE Plus Clinical resource.^22^ Three variants were in a disease gene inconsistent with the patient’s phenotype; and all these variants were classified as of uncertain significance by ACMG-AMP^23^ guidelines.

### Limitations and opportunities

Whilst our genotype-first method diagnosed patients missed by the initial 100KGP diagnostic strategy, it does not replace the importance of a phenotype-driven approach. It would be foolhardy not to look at all variants in a gene with a close phenotype match to the patient. Our method is best applied as a complementary screening strategy and will not diagnose the majority of patients, especially those with variants in non-constrained genes, or with pathogenic missense or extended splice site variants or with inherited variants. It also does not negate the need for variant curation, since some pLoF variants may not result in LoF.^24^

Our screening tool selects *de novo* variants, meaning that we excluded potential pathogenic variants in patients without trio data and excluded diseases whereby we may expect disease segregation e.g. cardiac or immune disease. Further, we selected genes in the first LOEUF decile to increase the specificity of our method. Expanding our approach to *de novo* pLoF variants in disease genes using a higher LOEUF threshold will inevitably increase the diagnostic yield, however this must be balanced with increased noise, and a greater number of biallelic disease genes. An alternative approach could be to use inheritance mode from OMIM plus a low LOEUF score to find haploinsufficient (versus gain of function) genes.

DeNovoLOEUF also risks identifying incidental findings including those in the ACMG secondary findings v3 list, which are a consequence of WGS and a tradeoff for increased diagnostic yield. Applying a LOEUF cut off <0.2 identified 10 genes in the ACMG list, although these genes could be manually removed from DeNovoLOEUF if preferred, or if the patient has not consented for secondary findings.^25^

Whilst DeNovoLOEUF has a high PPV, there are opportunities to refine our method to increase sensitivity at the expense of specificity. We propose a revised screening pipeline that will not only identify *de novo* variants in LOEUF constrained genes, but also screen for all known rare ClinVar pathogenic or likely pathogenic variants regardless of the gene panel applied (**Figure 4**). Our suggestion is to place *de novo* variants, ClinVar variants and novel coding variants into a new tier for assessment by NHS clinical laboratories to complement the current tiering system.^5^ Adding this additional review approach, along with a phenotype-driven variant analysis, is consistent with recently released best practices in genome analysis released from the Medical Genome Initiative.^26^ From reviewing 20 genomes, we estimate this approach would yield 3-9 potential additional variants per trio, using a LOEUF cut off <0.35, a missense constraint z-score >3,^27^ and likely pathogenic/pathogenic ClinVar entries with 2 stars or above. Our hope is to achieve a higher diagnostic yield per number of variants assessed by diagnostic labs, whereby we prioritise the most salient variants first.

**Figure 4.**
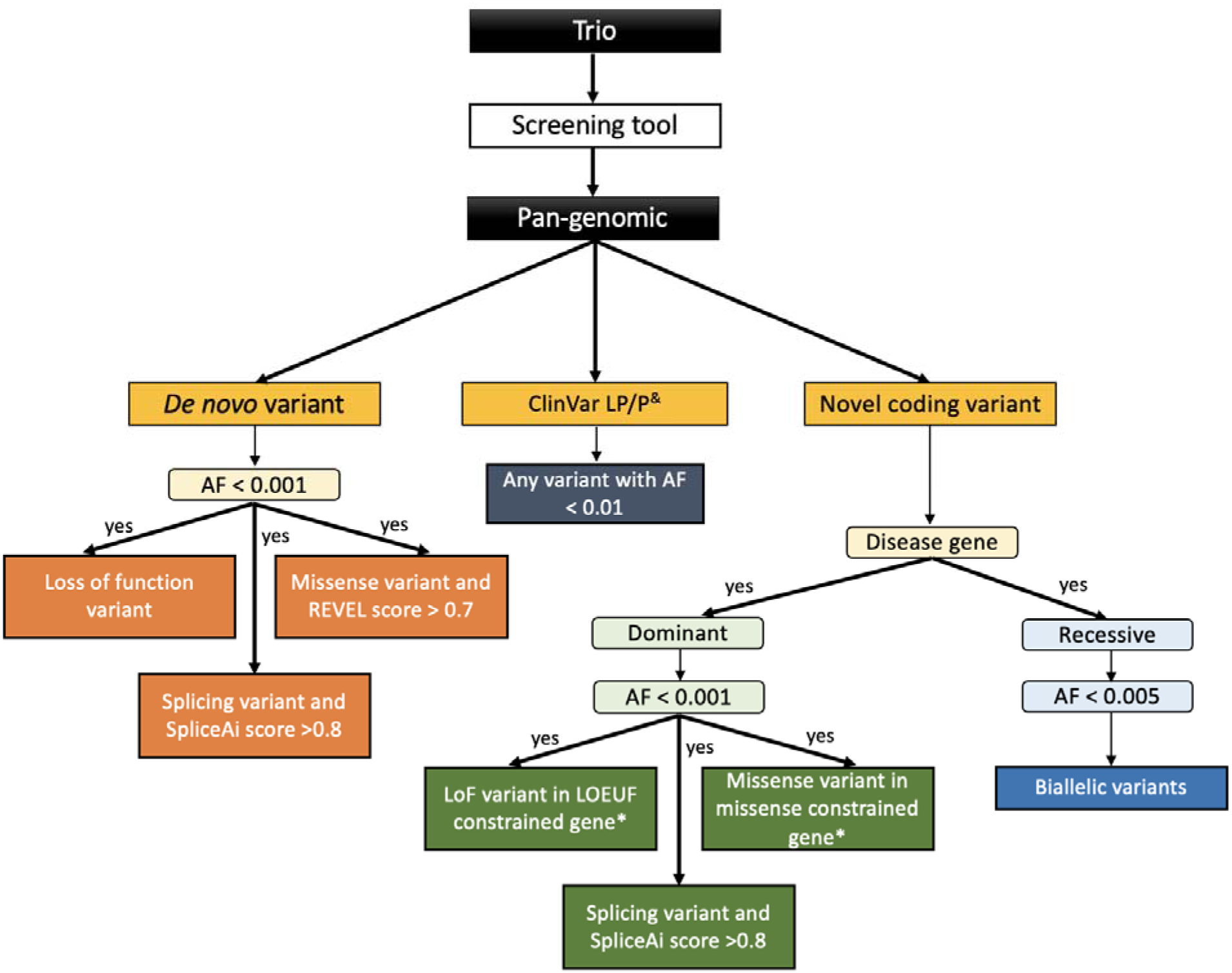
Proposed genotype-first screening strategy A possible refined screening strategy. All de novo variants with an allele frequency (AF) < 0.001 would be filtered, and loss of function variants would be prioritized, in addition to missense variants with a REVEL score >0.7, and splicing variants with a SpliceAI score > 0.8. All known ClinVar likely pathogenic (LP) and pathogenic (P) variants would be reviewed independent of zygosity and prioritized with an AF < 0.01. Novel coding variants (absent from population databases) in a known OMIM disease gene would be extracted and divided by disease mechanism. For dominant genes, variants are filtered with an AF of < 0.001. Retained variants would be prioritized if they were a loss of function (LoF) variant in a LOEUF constrained gene; a missense variant in a missense constrained gene, using a z-score >3 using the statistical method described by Samocha et al.^28^; or a splice site variant with a SpliceAI score >0.8. For recessive genes, variants are filtered with an AF < 0.005. Any biallelic (phased) variants are retained. ^*^Denotes user-specified cut offs for LOEUF and z-score missense constraint which can be tailored (or removed) as required. ^&^Suggest using ClinVar 2* and above.

## Conclusion

We present a targeted screening tool, DeNovoLOEUF, that can be applied at scale to rapidly identify putative pathogenic variants with a 96% positive predictive value. Our method complements current family-based analyses and can add value by identifying diagnostic variants missed by filtering strategies that adopt predefined disease-targeted gene panels. We have identified 39 pathogenic variants missed by the initial 100KGP variant prioritisation strategy. With 5 million more genomes being sequenced on the NHS, and many other international sequencing studies underway, we believe that our method alongside the new GEL initiative to report on Exomiser top ranked variants can help rapidly and effectively improve diagnostic efficiency and uplift diagnostic rates for the benefit of rare disease patients and their families.

## Data Availability

Access to the 100KGP dataset analysed in this study is only available as a registered GeCIP member in the Genomics England Research Environment, but restrictions apply to the availability of these data due to data protection and are not publicly available. Information regarding how to apply for data access is available at the following url: https://www.genomicsengland.co.uk/about-gecip/for-gecip-members/data-and-data-access/. All data shared in this manuscript were approved for export by Genomics England. The datasets and code supporting the current study have not been deposited in a public repository because the data are not public. Code showing data analysis on Genomics England data can be shared upon request within the Genomics England Research Environment.

## Supplementary information

Supplementary Data A: Excel spreadsheet of curated variants.

## Acknowledgements

This research was made possible through access to the data and findings generated by the 100,000 Genomes Project. The 100,000 Genomes Project is managed by Genomics England Limited (a wholly owned company of the Department of Health and Social Care). The 100,000 Genomes Project is funded by the National Institute for Health Research and NHS England. The Wellcome Trust, Cancer Research UK and the Medical Research Council have also funded research infrastructure. The 100,000 Genomes Project uses data provided by patients and collected by the National Health Service as part of their care and support. We would further like to extend our thanks to all the patients and their families for participation in the 100,000 Genomes Project. We are grateful to the Broad Center for Mendelian Genomics and Genome Aggregation Database teams for their helpful discussions in the development and application of constraint metrics in novel gene discovery.

